# Proteomics data in vitiligo: a scoping review

**DOI:** 10.1101/2024.02.26.24303359

**Authors:** Danique Berrevoet, Filip Van Nieuwerburgh, Dieter Deforce, Reinhart Speeckaert

## Abstract

An unbiased screening of which proteins are deregulated in vitiligo using proteomics can offer an enormous value. It could not only reveal robust biomarkers for detecting disease activity but can also identify which patients are most likely to respond to treatments. We performed a scoping review searching for all articles using proteomics in vitiligo. Eight manuscripts could be identified. Unfortunately, very limited overlap was found in the differentially expressed proteins between studies (15 out of 272; 5,51%) with variable degrees of the type of proteins and a substantial variety in the prevalence of acute phase proteins (range: 6-65%). Proteomics research has therefore brought little corroborating evidence on which proteins are differentially regulated between vitiligo patients and healthy controls or between active and stable vitiligo patients. While a limited patient size is an obvious weakness for several studies, an incomplete description of patient characteristics is an unfortunate and avoidable shortcoming. Additionally, the variations in the used methodology and analyses may further contribute to the overall observed variability. Nonetheless, more recent studies investigating the response to treatment seem to be more robust, as more differentially expressed proteins that have previously been confirmed to be involved in vitiligo were found. The further inclusion of proteomics analyses in clinical trials is recommended to increase insights into the pathogenic mechanisms in vitiligo and identify reliable biomarkers or promising drug targets. A harmonization in the study design, reporting and proteomics methodology could vastly improve the value of vitiligo proteomics research.

## 1. Introduction

Proteomics covers a variety of techniques for the comprehensive study of the structure, function, and expression of all proteins in a biological system, providing insights into cellular processes and molecular mechanisms at the protein level.(1) High-throughput proteomics can clinically be applied as a tool for large-scale characterization of differentially expressed proteins in relation to disease.(2) Therefore, it can play a key role in defining biomarkers in all different types of illnesses. Vitiligo is a pigmentary auto-immune disorder characterized by the destruction of melanocytes in the skin, giving rise to patchy depigmentation. This disease affects around 0.5-1% of the global population and can greatly impact the quality of life by increasing the risk of depression and anxiety.(3) Vitiligo is considered a multifactorial disease, including causes such as autoimmunity, neural dysregulation, oxidative stress and genetic predisposition.(4) To this day, the diagnosis of vitiligo continues to rely mainly on clinical presentation.(5) Current therapies to treat vitiligo include topical anti-inflammatory treatments, phototherapy, but also oral steroids, conventional immunosuppressants/immunomodulators, new targeted treatments such as Janus Kinase (JAK) inhibitors and surgical interventions.(6) However, these treatments are often only partially effective, cause adverse events and relapse of the disease can be seen after stopping treatment.(7) Therefore, the discovery of new biomarkers and treatment options is longed-for.(8,9)

A limited number of biomarkers has already been discovered for vitiligo through the discovery of the pathological role of the IFNγ-CXCL9/CXCL10-CXCR3 axis.(10) However, whereas many studies have focused on this IFNγ-pathway and on chemokines such as CXCL9/10, other important pathogenic factors might remain to be elucidated. Further research using an unbiased approach is required to obtain broader insights into the interplay of proteins such as interleukins (IL’s) and chemokines and their working mechanisms as potential therapeutic targets. Defining biomarkers through high-throughput proteomic analyses represents a promising approach for understanding the pathogenesis of vitiligo.(11) Interestingly, existing literature on proteomic studies in vitiligo reveals divergent results, emphasizing the necessity for a comprehensive overview of proteomic data in vitiligo.

The goal of this review was to answer the following question: “What proteins have been discovered in body fluids or whole tissue samples of vitiligo patients using high-throughput proteomic techniques?” The additional objective was to determine causes for possible different results in the included studies, and to see what research and experimental approaches could be implemented to achieve more reliable future proteomics studies.

## 2. Materials and Methods

A literature search was performed using both PubMed and Embase databases. ‘Proteomics studies’ were defined as studies using techniques that investigate a large number of proteins, not restricted to a limited predefined set of proteins which are believed to be relevant for the disease pathogenesis. All articles from inception to Nov 30^th^, 2023 were included. “Proteomics AND vitiligo” and “proteome AND vitiligo” were used as search queries to identify relevant studies. The systematic search was done by 2 independent researchers, DB and RS. Studies with the following objectives were included: Search for differentially expressed proteins between vitiligo patients and healthy controls (1) or between active and stable vitiligo patients (2). Additionally, studies that used high-throughput proteomics to monitor patients over time or to predict the response to treatments were considered. Reports with different aims were excluded. Only articles written in English and articles that investigated body fluids or whole tissue samples from vitiligo patients were considered. All original research studies using any type of study design were taken into account, including letters and abstracts. Both segmental and non-segmental vitiligo were considered. In case >1 DEPs list was reported for the same study population, only the most relevant DEPs list was included. Reviews or articles not presenting new proteomics data or publications using proteomics on individual cell types such as melanocytes, keratinocytes or immune cells were excluded. Additionally, patients with melanoma-associated leukoderma or other types of depigmentation were excluded. The study was conducted according to the PRISMA guidelines and the PRISMA flow chart can be found in the Supplementary Material (Supplementary Figure 1). The following data were extracted: differentially expressed proteins, study design, the description of patient characteristics, and the used proteomics technology. The overlap and difference between the obtained studies were analyzed. Subsequently, the literature was searched to determine if other studies confirmed the involvement of the top differentially expressed proteins that were reported in the included vitiligo studies. All differentially expressed proteins were categorized as immunologic or non-immunologic proteins, enzymes, hormones or proteases and labeled as acute-phase or non-acute-phase proteins by DB and RS based on the literature. To optimize the readability of this review, the results section also includes aspects of interpretability.

## 3. Results

The initial search yielded 72 records from the two databases, of which 51 remained after the removal of duplicates. Finally, eight records were included after abstract and full-text screening, and eligibility assessment.

### Studies investigating vitiligo patients versus healthy controls or active (i.e. progressive) versus stable patients

Very divergent results were reported for the differentially expressed proteins in vitiligo with limited overlap between publications. An overview of the described differentially expressed proteins and methods in this review can be found in Table 1. In the 5 studies that compared vitiligo patients with healthy controls or active versus stable patients, 272 differentially expressed proteins were found of which 15 (5.51%) were detected in more than one study (Table 2 & 3). For only 7 (2.57%) of these proteins it concerned a publication of different research groups as one group published 2 studies.

**Table 1:**
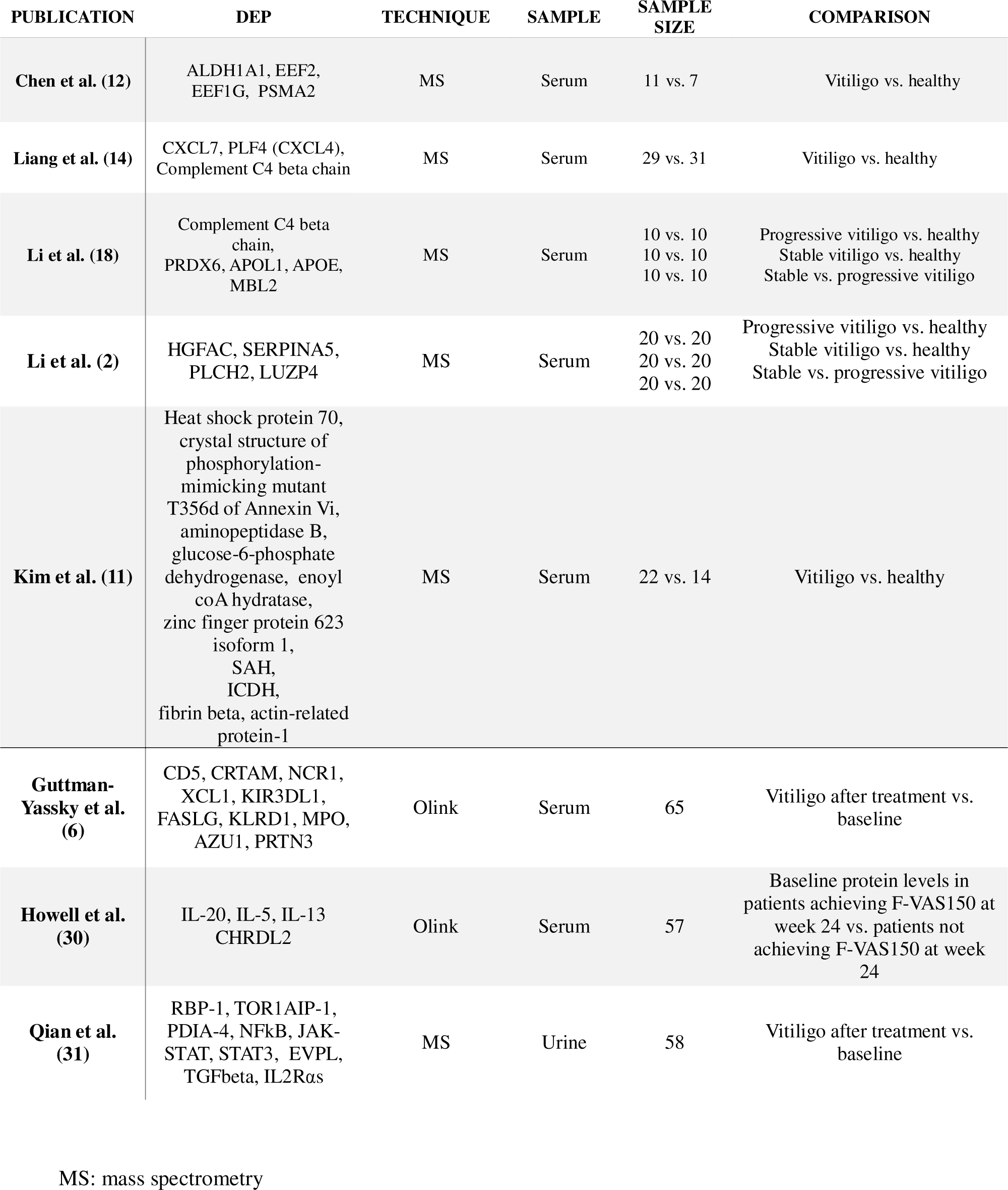
Essential results and methods used in the vitiligo proteomics studies.

**Table 2:** Similarities in differentially expressed proteins in different proteomics studies – vitiligo patients versus healthy controls.

| PROTEIN | IDENTIFICATION OF THE STUDY |
| --- | --- |
| Apolipoprotein A-I | Li et al. (Feb 2018) (2)<br>Li et al. (Feb 2018) (18) |
| Cathepsin D | Li et al. (Feb 2018) (2)<br>Liang et al. (Oct 2019) (14) |
| Complement C4-B | Li et al. (Feb 2018) (2)<br>Liang et al. (Oct 2019) (14)<br>Li et al. (Feb 2018) (18) |
| Complement C4-A | Li et al. (Feb 2018) (2)<br>Liang et al. (Oct 2019) (14)<br>Li et al. (Feb 2018) (18) |
| Haptoglobin | Li et al. (Feb 2018) (2)<br>Li et al. (Feb 2018) (18) |
| Haptoglobin-related protein | Li et al. (Feb 2018) (2)<br>Chen et al. (May 2023) (12) |
| Ig alpha-2 chain C region | Li et al. (Feb 2018) (2)<br>Li et al. (Feb 2018) (18) |
| Ig gamma-4 chain C region | Li et al. (Feb 2018) (2)<br>Li et al. (Feb 2018) (18) |
| Ig mu chain C region | Li et al. (Feb 2018) (2)<br>Li et al. (Feb 2018) (18) |
| Inter-alpha-trypsin inhibitor heavy chain H4 | Li et al. (Feb 2018) (2)<br>Li et al. (Feb 2018) (18) |
| Keratin, type II cytoskeletal 1 | Li et al. (Feb 2018) (2)<br>Li et al. (Feb 2018) (18) |
| Platelet factor 4 | Li et al. (Feb 2018) (2)<br>Liang et al. (Oct 2019) (14) |
| Thrombospondin-1 | Liang et al. (Oct 2019) (2)<br>Li et al. (Feb 2018) (14) |
| Vitamin D-binding protein | Li et al. (Feb 2018) (2)<br>Liang et al. (Oct 2019) (14) |

A study by Chen et al. carried out a proteomic analysis using mass spectrometry on serum of 11 patients of the Chinese Han population with active vitiligo versus 7 healthy controls, validated by another 10 vitiligo patients and 10 healthy controls. The acquired raw data was analyzed using Proteome Discoverer (version 2.4). The mean extent was 5.55% in the proteomics group and 6.18% in the validation group. Active disease was defined as no new lesions or enlargement of existing lesions in the last 6 months [Vitiligo Disease Activity score (VIDA) ≥ 2]. Thirty-one differentially expressed proteins (DEPs) (P<0.05, fold change >1.2) were found out of 1019 proteins (3.04%), of which 21 were upregulated, and 10 were downregulated. Aldehyde dehydrogenase 1A1 (ALDH1A1), elongation factor 2 (EEF2), proteasome subunit alpha type-2 (PSMA2) and elongation factor 1-gamma (EEF1G) showed to be the upregulated proteins with the highest fold change level compared to 7 healthy controls.(12) ALDH1A1 is an enzyme that converts lipid aldehydes to lipid carboxylic acids. Research has shown that it potentially regulates the melanogenesis by converting 9-cis retinal to retinoic acid, identified as a pigment stimulatory agent.(13) The role of EEF2, EEF1G and PSMA2 in vitiligo or inflammation pathways remains unclear.

The research group of Liang et al. also performed a proteomic analysis on serum using mass spectrometry. The data analysis of this study was performed with the Proteome Discoverer software (version 1.6). Here, 78 DEPs were found out of 582 proteins (13.40%), when comparing 29 non-segmental vitiligo patients to 31 healthy controls of whom sex and age were matched. However, essential patient characteristics such as disease activity and disease extent were not specified. Additionally, differentially expressed proteins were defined by a fold-change of >1.5. Statistical significance was not specified. This research group reported that many proteins involved in the immune system, such as CXCL7, PLF4 (CXCL4) and the complement C4 beta chain were elevated in serum samples of vitiligo patients compared to the control group.(14) CXCL4 is released by platelets and involved in atherosclerosis.(15) CXCL4 is an agonist of CCR1 and may play a role in inflammation by driving monocyte migration. Elevated values of CXCL4 have not been confirmed by other studies in vitiligo. Nonetheless, increased CXCL4 levels would be consistent with an upregulated CXCR3 signaling in vitiligo.(16) CXCL7 is also released from activated platelets and orchestrates neutrophil recruitment. Vitiligo is however not a neutrophilic disorder leaving the possible implication of CXCL7 largely unexplained.(17) Interestingly, CXCL10 could not be detected in this study, despite it already being reported in numerous publications to be significantly elevated in vitiligo patients compared to healthy controls. According to the authors of this study, this might be attributed to its potentially low levels in serum, although the lack of information about disease activity in these patients makes this difficult to assess. CXCL10 has been detected by many studies in the serum of vitiligo patients with expression levels well above the detection limits of standard ELISA kits and multiplex bead arrays.(8)

In a study executed by Li et al., the serum of 10 patients with progressive vitiligo and 10 patients with stable vitiligo were compared to 10 healthy controls and to each other.(18) Data analysis was performed using Statistical Package for Science Software (SPSS) version 16.0. Progressive vitiligo was defined as the enlargement of original lesions or the occurrence of new depigmentation from 6 weeks to a year, while stable vitiligo was defined as lesions being stable for more than 1 year or with spontaneous repigmentation. No data were provided on the vitiligo extent. Forty-eight DEPs were further analyzed by mass spectrometry after two-dimensional gel electrophoresis. Statistical significance was attributed to differences with P-values below 0.05, and protein spots exhibiting an expression difference of at least 2-fold were classified as differentially expressed. The total amount of acquired spots on the gel was not reported. Most identified proteins had an enzyme regulator activity, followed by ion binding, peptidase activity, lipid binding, and enzyme binding. Complement C4-B was also reported as a DEP in this publication, however displaying contradicting results in the different groups. Verification of protein expression was performed by western blotting. This analysis showed peroxiredoxin-6 (PRDX6) and apolipoprotein L1 (APOL1) to be downregulated in patients with progressive and stable vitiligo compared to healthy controls, with in both cases the protein being more downregulated in progressive vitiligo compared to stable vitiligo. PRDX6 has been reported as an activator of inflammatory pathways, as well as a protective mechanism by counteracting increased reactive oxygen species and repairing the membranes of oxidized cells caused by oxidative stress.(19) APOL1 has shown to be upregulated in response to inflammation through the Janus kinase signal transducer and activator of transcription (JAK-STAT) pathway, a pathway also involved in the vitiligo pathogenesis.(20) Additionally, apolipoprotein E (APOE) was also reported to be downregulated in progressive as well as stable vitiligo patients compared to healthy controls, with the protein being more downregulated in stable vitiligo than the progressive form. Different sources imply that this protein has anti-inflammatory as well as pro-inflammatory properties.(21–23) Finally, mannose-binding protein C (MBL2) was downregulated in progressive vitiligo compared to healthy controls, while it was upregulated in stable vitiligo patients compared to controls. This protein has been reported as an acute phase protein in response to inflammation. However, this statement has been the subject of extensive debate, due to its heterogeneous properties.(24)

Another study executed by this research group performed a serum analysis of patients with stable (n=20) or progressive vitiligo (n=20), and of healthy controls (n=20). Data analysis was performed using the protein pilot software 5.0. Patients with stable lesions for at least a year were considered stable. Progressive vitiligo (VIDA ≥ 2) was defined by the occurrence of new skin lesions, the expansion of original skin lesions or the development of the Koebner phenomenon within three months. Again, no information on disease extent was provided. Using mass spectrometry 171 differentially expressed proteins were identified (80 in stable patients and 89 in progressive patients, compared to healthy controls). The total amount of detected proteins was not reported. Additionally, differentially expressed proteins were not specified by fold-change or statistical significance. Seventy-one DEPs were found between active versus stable patients. The highest upregulation was observed for the proteins HGFAC and SERPINA5 in both stages compared to the healthy control group.(2) Hepatocyte growth factor activator (HGFAC) is a member of the peptidase family S1 and is a serine protease converting hepatocyte growth factor to its active form.(25) SERPINA5, also called Protein C inhibitor, limits the activity of the anticoagulant protein C. It acts as an anti-inflammatory factor in severe inflammatory disorders, such as sepsis.(26) Phospholipase C Eta 2 (PLCH2) expression was upregulated in stable vitiligo patients, and significant downregulation of LUZP4 in progressive vitiligo patients was seen. The mechanisms of how PLCH2 or leucine zipper protein 4 (LUZP4) could be involved in inflammation and vitiligo remains unclear. To date, confirmatory data are missing for all 4 identified proteins.

A completely different set of DEPs were detected when applying high-throughput proteomics on the serum of 22 patients with non-segmental generalized active vitiligo with unclear vitiligo extent compared to 14 healthy controls in a study by Kim et al.(11) Active vitiligo was defined by the enlargement or reoccurrence of depigmentation within 3 months. A total of around 2000 protein spots were observed after two-dimensional gel electrophoresis. Ten (0.5%) of these were not observed in the sera of control subjects and were therefore studied further by mass spectrometry. Data analysis was performed using the MS-Fit program provided by UCSF (NCBI scanning algorithm). Differentially expressed proteins were again not specified by fold-change or statistical significance. Heat shock protein 70, aminopeptidase B, glucose-6-phosphate dehydrogenase, zinc finger protein 623 isoform 1, S-adenosylhomocysteine hydrolase (SAH), actin-related protein 1, NADP-dependent isocitrate dehydrogenase (ICDH), crystal structure of phosphorylation-mimicking mutant T356d of Annexin Vi, enoyl coA hydratase, and fibrin beta were reported to be upregulated compared to the healthy control group. Increased heat shock protein 70 levels have been reported in the skin of vitiligo patients and linked with disease progression.(27) Heat shock protein 70 activates plasmacytoid dendritic cells leading to the production of IFN-α. This in turn induces CXCL9 and CXCL10 expression by keratinocytes.(28) Glucose-6-phosphate dehydrogenase levels have been repeatedly reported to be decreased in vitiligo.(29) Remarkably, the other proteins have not been identified as significant differentially expressed proteins in any subsequent publications where proteomics techniques were employed.

### Studies investigating the response to treatment

Proteomics was used in a recent study by Guttman-Yassky et al. to investigate the effect of ritlecitinib, a JAK3/TEC family kinase inhibitor, on the protein levels of 65 non-segmental vitiligo patients.(6) The patients had a mean involved body area of 16.8% and all patients had active disease at baseline. Serum aliquots were analyzed using the Olink Proseek multiplex assay, using the inflammation, cardiovascular disease I/II and neurology panels. Here, a decrease was shown for cluster of differentiation 5 (CD5), Cytotoxic And Regulatory T Cell Molecule (CRTAM), Natural cytotoxicity triggering receptor 1 (NCR1), X-C motif chemokine ligand 1 (XCL1), Killer cell immunoglobulin-like receptor 3DL1 (KIR3DL1), Fas ligand (FASLG) and killer cell lectin like receptor D1 (KLRD1) when applying ritlecitinib after 4 and 24 weeks compared to baseline and placebo controls (n=14). All these biomarkers are involved in T cell/T cell activation/NK activation. A significant downregulation was also seen for myeloperoxidase (MPO), azurocidin (AZU) and proteinase 3 (PRTN3), which are all atherosclerosis biomarkers.(6)

Howell et al. performed proteomics (Olink Proximity extension assay using the oncology II, cardiovascular disease II, cardiovascular disease III, inflammation I, neurology I, immune response, metabolism, organ damage, cardiometabolic, cell regulation, development, and neuroexploratory panels) on serum at baseline to identify biomarkers for repigmentation due to ruxolitinib.(30) All 57 patients suffered from extensive vitiligo (total BSA: 20.54%-28.53%) with half of the patients having active disease. Seventy-six of the 1104 (6,88%) tested proteins were differentially expressed.(30) Patients with ≥50% improvement in facial Vitiligo Area Scoring Index scores (F-VASI50) by 24 weeks displayed 10 upregulated proteins, whereas patients with less than 50% improvement in the face carried 64 elevated proteins. Remarkably, many of the increased proteins in patients achieving F-VASI50 were interleukins such as IL-20 and Th2 linked cytokines IL-5 and IL-13 for which no major role in vitiligo has previously been described. CHRDL2 was also increased and associates with members of the transforming growth factor beta superfamily. In patients not achieving F-VASI50, CXCL9 and CXCL10 were the 5^th^ and 15^th^ most upregulating proteins which are both highly linked to vitiligo activity in numerous publications.(16)

Qian et al. used a different approach by using urine samples to predict the efficacy of corticosteroid treatment and monitor the disease.(31) Forty-two patients were classified as good responders, while 16 patients were considered to be resistant to treatment. Using mass spectrometry, 245 and 341 differentially expressed proteins were found between both groups before and after corticosteroid treatment, respectively. Further ELISA analysis was done to validate the changed proteins. Retinol binding protein-1 (RBP-1), torsin 1A interacting protein 1 (TOR1AIP-1), and protein disulfide isomerase family A member 4 (PDIA-4) were considered as potential markers for treatment success. Only RBP-1 has previously been reported to be involved in glucocorticosteroid-related signaling.(32,33) Several pathways involved in corticosteroids resistance were enriched. Additionally, changes in many immunological signals were found including the nuclear factor kappa-light-chain-enhancer of activated B cells (NFkB), Janus kinase/signal transduction (JAK-STAT), signal transducer and activator of transcription 3 (STAT3), evoplaking (EVPL), transforming growth factor beta (TGFbeta) signaling, and IL2Rαs of which several have been confirmed to be implicated in vitiligo.(9,31,34)

### Comparison of the function of the DEPs

When comparing the identified DEPs between vitiligo patients and healthy controls (Figure 1), a predominant result (53%) of immunologic DEPs (of which 46% are proteins, 5% enzymes, 1% proteases and 1% hormones) can be observed. The DEPs were considered immunological if they have been described as prominent players in inflammatory pathways. When looking at the individual studies, only the papers published by Li et al. show a majority of immunologic DEPs.(2,18) One of these two studies reports the largest number of DEPs (125 DEPs compared to 33; 78; 26; 10 DEPs in the other studies), therefore contributing to the power of this result. The DEPs determined in the studies investigating response to treatment (Figure 2), show a predominant result (59%) of non-immunologic DEPs (of which 39% are proteins,19% enzymes and 1% proteases). Nevertheless, when again looking at the individual studies, the studies performed by Guttman et al. and Howell et al. show a majority of immunologic DEPs.(6,30)

**Figure 1:**
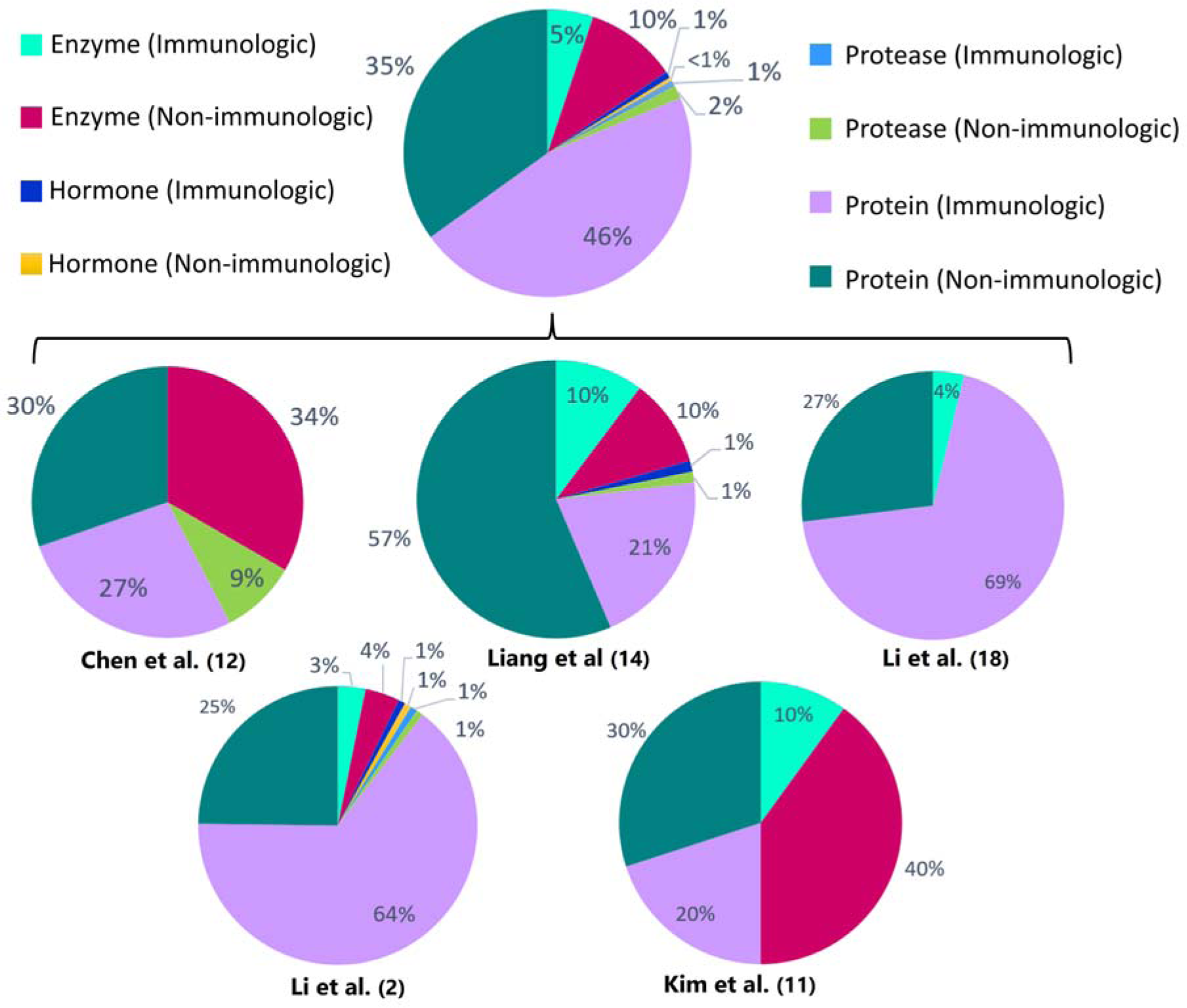
Percentages of immunologic and non-immunologic proteins, enzymes, hormones and proteases combined and in the individual different proteomics studies investigating vitiligo patients versus healthy controls or active versus stable patients. Enzymes are defined as all enzymes that are not proteases. Proteins are defined as proteins that can’t be classified into the other depicted groups (enzymes, proteases, hormones). The proteins are considered immunologic if they have been described as prominent players in inflammatory pathways.

**Figure 2:**
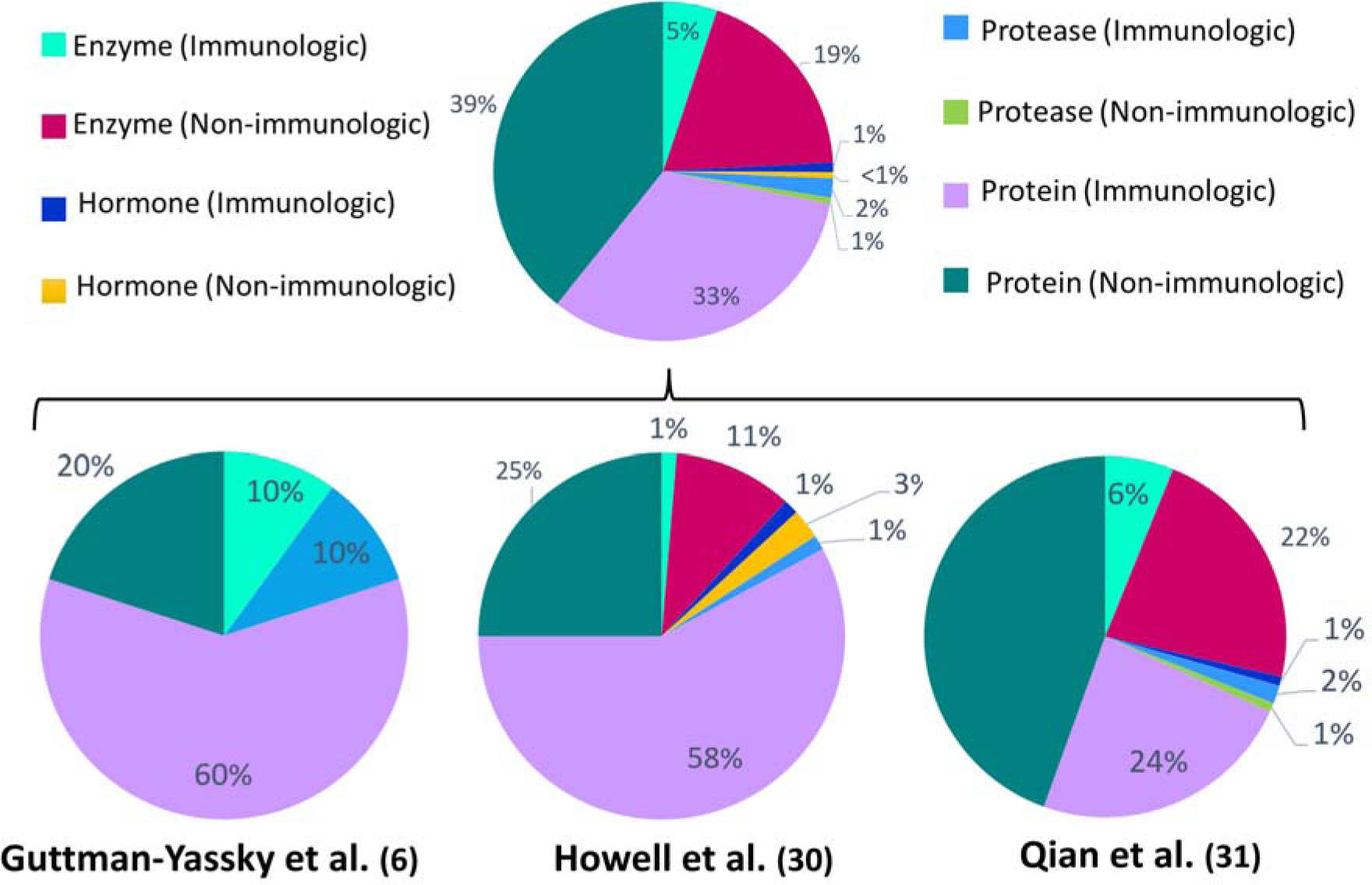
Percentages of immunologic and non-immunologic proteins, enzymes, hormones and proteases combined and in the individual different proteomics studies investigating response to treatment. Enzymes are defined as all enzymes that are not proteases. Proteins are defined as proteins that can’t be classified into the other depicted groups (enzymes, proteases, hormones). The proteins are considered immunological if they have been described as prominent players in inflammatory pathways.

In all studies, except for one performed by Li et al., the identified DEPs are mainly non-acute phase proteins, meaning they are not elevated or decreased during inflammation. Nonetheless, a broad range in the presence of acute phase proteins was observed between studies, ranging between 6% and 65%. This suggests that inconsistent differences in inflammatory status between comparison groups influences the detection of DEPs (Figure 3). Acute phase proteins were defined as proteins that are elevated or lowered during inflammation. Of the 15 overlapping DEPs across the 5 publications depicted in Tables 2 and 3, 13 DEPs (86,67%) are defined as immunological proteins, indicating their prominent role in inflammatory pathways. Additionally, 7 (46,67%) out of these 15 DEPs are acute phase proteins. These findings add to the understanding that inflammatory pathways play a significant role in the pathogenesis of vitiligo. To determine if these overlapping DEPs are potential drug targets, further verification of their levels in vitiligo patients and identification of their role in this pathogenesis is required.

**Figure 3:**
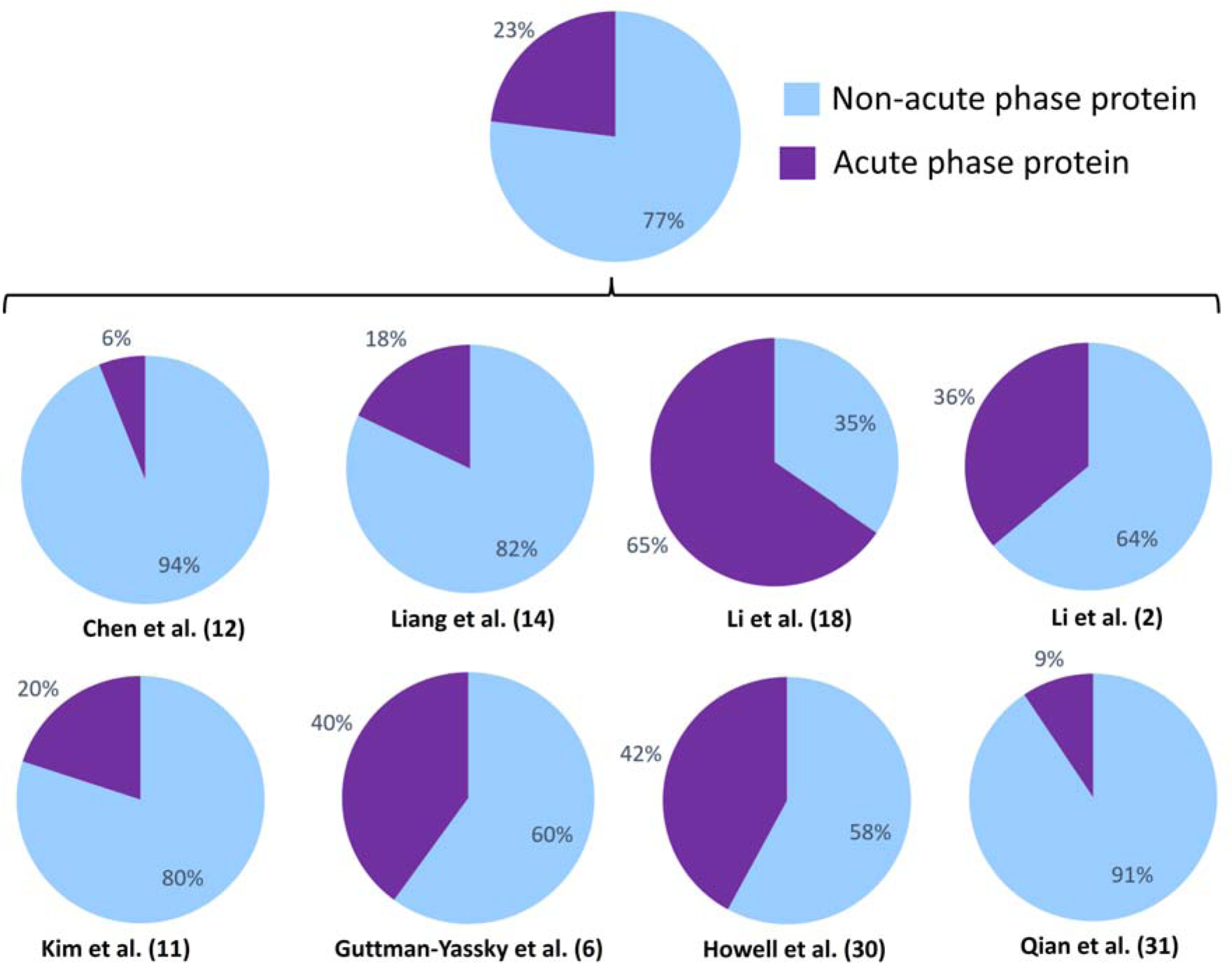
Percentages of acute phase proteins combined and in the individual different proteomics studies. Acute phase proteins are defined as proteins that are elevated or lowered during inflammation.

**Table 3:** Similarities in differentially expressed proteins in different proteomics studies – stable versus active vitiligo.

| PROTEIN | IDENTIFICATION OF THE STUDY |
| --- | --- |
| Complement C4-A | Li et al. (Feb 2018) (2)<br>Li et al. (Feb 2018) (18) |
| Complement C4-B | Li et al. (Feb 2018) (2)<br>Li et al. (Feb 2018) (18) |
| Ig mu chain C region | Li et al. (Feb 2018) (2)<br>Li et al. (Feb 2018) (18) |
| Immunoglobulin J chain | Li et al. (Feb 2018) (2)<br>Li et al. (Feb 2018)(18) |

## 4. Discussion

An unbiased detection of proteins that are upregulated or downregulated in vitiligo holds huge promise to learn more about the main driving factors that could entail biomarkers or promising treatment targets. However, the characterization of proteomic data in vitiligo poses a significant challenge due to the pronounced divergence observed across published studies. Discrepancies arise from variations in experimental methodologies, sample populations, analytical platforms and different research questions. Multiple investigations employing high-throughput proteomic techniques have reported incongruent results, hindering the establishment of a unified understanding of the protein landscape in vitiligo. Sample sizes in all discussed studies were rather small, reducing statistical power when investigating a great number of proteins. Additionally, patient characteristics are often incompletely described. The disease extent of the included patients is even not mentioned in 4 out of 5 studies investigating vitiligo patients versus healthy controls or active versus stable patients. A description of the treatment of patients is also missing in several publications. Additionally, the definitions assigned to ‘stable’ or ‘active’ patients lack uniformity across the diverse publications. All these limitations drastically decrease the interpretability of these reports with uncertain relevance of the reported results. Given the limited overlap between differentially regulated proteins in the different studies it is impossible to extract clear conclusions leading to more insights for vitiligo.

Many of the common DEPs across publications, documented in Tables 2 and 3, originated from the research team of Li et al., albeit in distinct publications.(2,18) This prompts us to contemplate whether this occurrence is mainly attributed to this research group employing their own standard techniques and data analysis programs across these two publications. Additionally, the research group of Li et al. was the only research group included in this review that compared active to stable vitiligo patients in both of their publications. Thus, the results of Table 2 only illustrating overlapping DEPs between the two studies of the same research group is not due to a lack of these proteins in other studies, but rather to the fact that only these studies could be compared with each other.

Studies using proteomics to determine the response to treatment or using repeated samples to follow the evolution during treatment are more robust and seem indeed to detect more proteins of which the involvement in vitiligo has been previously demonstrated. The decrease of biomarkers involved in T and NK cell activation in patients receiving ritlecitinib is highly likely to represent the working mechanisms of this JAK3/TEC inhibitor.(6) The results of Howell et al. performed at baseline to predict repigmentation due to ruxolitinib also identified several chemokines which are known to be increased in vitiligo.(30) This induces confidence in the validity of the obtained results.

The included study by Howell et al. is a post-hoc analysis of a study conducted by Rosmarin et al., using the same proteomics analysis and patient population.(30,35) In the study by Rosmarin et al, DEPs are described between baseline and 24 weeks of treatment (ruxolitinib), without taking the treatment outcome (F-VASI50) into account. However, this study identified a small number of DEPs that were not reported by Howell et al., but were described by other studies included in this review. These include carbonic anhydrase 4 and ephrin A4, described by Qian et al., and CRTAM, NCR1, XCL1 and FASLG, reported by Guttman et al.(6,31)

The importance of the inflammatory pathways in the pathological mechanism of vitiligo is already extensively described in numerous scientific papers. Therefore, we would expect to detect a majority of chemokines (e.g. CXCL10, CXCL11) and cytokines (e.g. IL-17, IL-23) being differentially expressed across the different publications when comparing vitiligo patients to healthy controls or active vs stable vitiligo patients. However, this seems not to be the case. This can be explained by the technical aspects of the experiments employed in these publications. In this review, we specifically looked for DEPs identified by high-throughput proteomics techniques. In 6 out of the 8 included publications, mass spectrometry was used, although using different types of ionization and detection techniques. Due to its low sensitivity, this technique is unfavorable to detect proteins such as cytokines and chemokines, as these are low abundant proteins. More sensitive, targeted assays are required to obtain this, such as the ELISA assay or by using targeted panels.

Furthermore, findings obtained by mass spectrometry can differ depending on instrumental variability, diverse sample preparation or experimental conditions and settings, explaining the wide variety of inconsistent outcomes. Addressing and understanding these factors is essential for obtaining reliable and reproducible results in mass spectrometry experiments. Validation experiments and quality control measures are often performed to minimize variability and ensure the accuracy of their findings.(36) Divergent findings extend to the identification and quantification of key proteins implicated in vitiligo, such as those involved in immune response modulation and melanocyte function. This lack of consensus underscores the necessity for methodological standardization and harmonization of experimental procedures within the field of vitiligo proteomics. The incorporation of stringent quality control measures and the establishment of reference datasets could enhance result reproducibility and comparability across studies.

Moreover, understanding the precise changes in protein composition over time at various phases of vitiligo progression is crucial for unraveling the complex molecular mechanisms underlying the illness. Performing high-throughput proteomics on lesional skin cells instead of serum, could provide us with more specific insights into the local molecular changes associated with the condition, allowing for a more localized understanding of the underlying mechanisms and potential therapeutic targets. Integrative approaches, such as merging proteomic data with complementary omics datasets, could also provide a more comprehensive understanding of the complex interplay between genetic, transcriptomic, and proteomic factors in vitiligo etiology. Collaborative efforts toward standardization and the establishment of shared resources within the scientific community are necessary to navigate and resolve the current challenges associated with the heterogeneous nature of vitiligo proteomic research. Additionally, emphasis should be placed on the validation of discovered biomarkers and integrating proteomic findings with other omics data, such as genomics and metabolomics, for a comprehensive understanding of the disease.(37,38)

## 5. Conclusion

The current state of proteomics research in vitiligo seems not to have reached its full potential. Several studies are hampered by a limited number of patients, incomplete description of patient characteristics and differences in the used methodology and analyses. The limited overlap in differentially expressed proteins across the different studies is concerning, and drawing substantiated conclusions about the involved DEPs in vitiligo with a satisfactory level of certainty is therefore unattainable. However, studies using proteomics to assess response to treatment and especially designs with repeated blood sampling are highly valuable and offer interesting insights. Maximizing the great potential of proteomics to further enhance our understanding of the pathogenic mechanisms in vitiligo can be achieved through the integration of unified clinical data reporting, harmonized proteomics methodology, and standardized study designs.

## Data Availability

All data produced in the present work are contained in the manuscript

