## Supplementary figures and images for "Proteomics data in vitiligo: a scoping review"

### Supplementary Figure 1

**Supplementary Figure 1: PRISMA Flow Chart**

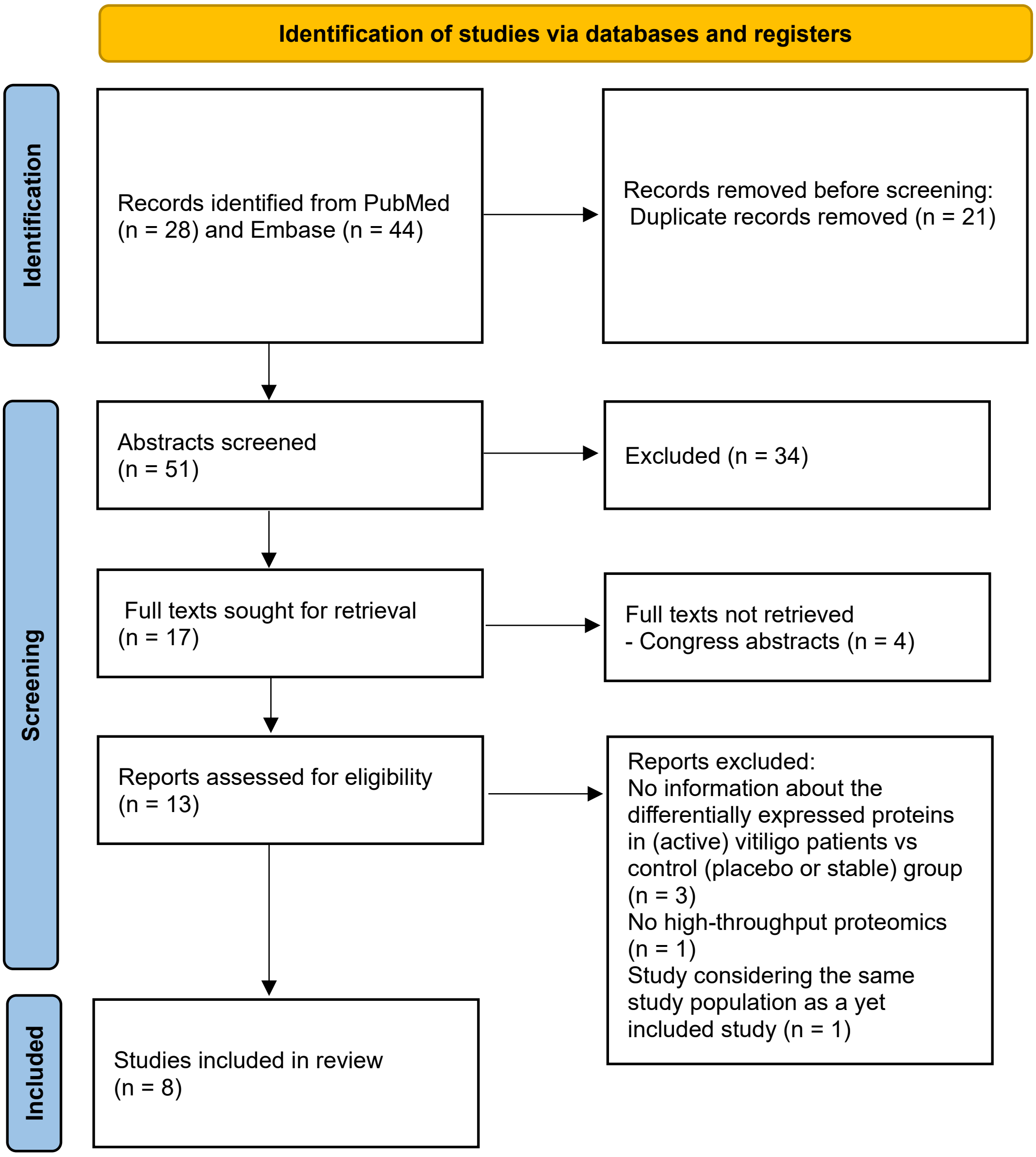
